# A SARS-CoV-2 variant of concern triggers Fc effector function with increased cross-reactivity

**DOI:** 10.1101/2021.11.05.21265853

**Authors:** Simone I. Richardson, Nelia P. Manamela, Boitumelo M. Motsoeneng, Haajira Kaldine, Frances Ayres, Zanele Makhado, Mathilda Mennen, Sango Skelem, Noleen Williams, Nancy J. Sullivan, John Misasi, Glenda G. Gray, Linda-Gail Bekker, Veronica Ueckermann, Theresa M. Rossouw, Michael T. Boswell, Ntobeko A. B. Ntusi, Wendy A. Burgers, Penny L. Moore

**Affiliations:** National Institute for Communicable Diseases of the National Health Laboratory Services, Johannesburg, South Africa; MRC Antibody Immunity Research Unit, School of Pathology, University of the Witwatersrand, Johannesburg, South Africa; Vaccine Research Center, National Institute of Allergy and Infectious Diseases, National Institutes of Health, Bethesda, MD 20892, USA; The South African Medical Research Council, Tygerberg, South Africa; The Desmond Tutu HIV Centre, University of Cape Town, Cape Town, South Africa; Division for Infectious Diseases, Department of Internal Medicine, Steve Biko Academic Hospital and University of Pretoria, Pretoria, South Africa; Department of Immunology, Faculty of Health Sciences, University of Pretoria, Pretoria, South Africa; Institute of Infectious Disease and Molecular Medicine, University of Cape Town, Cape Town, South Africa; Cape Heart Institute, Faculty of Health Sciences, University of Cape Town, Cape Town, South Africa; Department of Medicine, University of Cape Town and Groote Schuur Hospital, Cape Town, South Africa; Division of Medical Virology, Department of Pathology, University of Cape Town, Cape Town, South Africa; Wellcome Centre for Infectious Diseases Research in Africa, University of Cape Town, Cape Town, South Africa; Centre for the AIDS Programme of Research in South Africa, Durban, South Africa

**Author notes:** **Corresponding author:** Penny L. Moore.

**Keywords:** Variant of concern, SARS-CoV-2, Fc effector function, Ad26.COV2.S, Beta, Delta

## Abstract

SARS-CoV-2 variants of concern (VOCs) exhibit escape from neutralizing antibodies, causing concern about vaccine effectiveness. However, while non-neutralizing cytotoxic functions of antibodies are associated with decreased disease severity and vaccine protection, Fc effector function escape from VOCs is poorly defined. Furthermore, whether VOCs trigger Fc functions with altered specificity, as has been reported for neutralization, is unknown. Here, we demonstrate that the Beta VOC partially evades Fc effector activity in individuals infected with the original (D614G) variant. However, not all functions are equivalently affected, suggesting differential targeting by antibodies mediating distinct Fc functions. Furthermore, Beta infection triggered responses with significantly improved Fc cross-reactivity against global VOCs compared to either D614G infected or Ad26.COV2.S vaccinated individuals. This suggests that, as for neutralization, the infecting spike sequence impacts Fc effector function. These data have important implications for vaccine strategies that incorporate VOCs, suggesting these may induce broader Fc effector responses.

## Introduction

Continued SARS-CoV-2 transmission worldwide in the absence of adequate vaccine coverage has resulted in the emergence of several viral variants of concern (VOCs) including Alpha (B.1.1.7), Beta (B.1.351/501Y.V2), Gamma (P.1) and Delta (B.1.617.2). These VOCs are generally able to evade neutralizing responses, with Beta, first identified in South Africa in October 2020^1^, showing the greatest capacity for neutralization escape from convalescent and vaccinee sera ^2–6^. In contrast, preservation of T cell function and spike binding antibody levels has been described for several VOCs after both infection and vaccination ^4,7–9^. In addition to mediating neutralization, antibodies drive several effector functions through their ability to engage cellular receptors via their Fc portion, including antibody-dependent cellular cytotoxicity (ADCC), cellular phagocytosis (ADCP), cellular trogocytosis (ADCT) or cell membrane nibbling and complement deposition (ADCD). Therefore, the presence of cross-reactive binding antibodies is consistent with reports of preserved Fc effector function in convalescent sera and after vaccination, and with effectiveness data showing that several vaccines maintain effectiveness against VOCs ^2,7,10^. For example, the Ad26.COV.2S vaccine maintained efficacy against severe COVID-19 illness as Beta emerged in South Africa despite substantially reduced neutralization titers^2,4,5,11^.

The majority of antibodies elicited in response to infection are non-neutralizing ^12^. As mutations in VOCs occur primarily in the RBD and the N-terminal domains targeted by neutralizing antibodies, antibodies able to bind outside of these sites and mediate potent antiviral function may be crucial for protection from severe infection. As for many other diseases, Fc effector function has been associated with reduced COVID-19 severity and mortality, suggesting an important early role for these functions in disease outcomes^13,14^. Furthermore, isolated antibodies from convalescent donors require Fc function for optimal protection and therapeutic efficacy ^15,16^. Fc functions have also been shown to persist beyond neutralizing responses following SARS-CoV-2 infection, suggesting that these may be an important target for vaccine design ^17,18^.

Fc effector function correlated with protection through vaccination in non-human primates ^10,19,20^and is potently elicited by vaccination in humans ^2,7,21,22^. Beyond this, nuances in the magnitude and breadth of Fc receptor binding responses from convalescent donors and different vaccine regimens suggests that these responses vary in response to stimulation by specific antigens, formulations or doses ^23^.

For neutralization, there is strong evidence that the sequence of the infecting virus impacts the breadth of the resulting neutralizing antibodies ^3,9,24^. Neutralizing antibodies triggered by VOCs show varying patterns of breadth compared to the original D614G and one another, suggesting that spikes with different genotypes differentially impact the repertoire of antibodies that they triggered. However, similar studies characterizing Fc effector functions in individuals infected with variants of concern have not yet been conducted. Since March 2020, South Africa has experienced three distinct waves of COVID-19 infection, each dominated by a different variant. In this study we leveraged these virologically distinct waves to define both Fc effector response escape from VOCs, and to describe Fc responses to VOCs. We used convalescent sera from individuals infected with Wuhan D614G to show that Beta partially evades several Fc effector functions. However, individuals infected with Beta developed Fc effector function with improved cross reactivity for all VOCs. Lastly, we show that Fc effector function elicited by the Ad26.COV.2.S vaccine is largely retained across VOCs, but is not as cross-reactive as those elicited by Beta. These findings illustrate that VOCs differentially trigger Fc effector functions, with important implications for vaccination.

## Results

### Binding antibodies, unlike neutralizing antibodies, retain high level activity against heterologous variants

South Africa’s first wave peaked in mid-July 2020, the second in January 2021 and the third in August 2021 (Figure 1A). The first wave was dominated by original Wuhan D614G, the second wave was driven by the Beta variant, and the third wave was almost exclusively Delta. We made use of convalescent plasma from the first two waves (first wave n = 27; second wave n = 21) to determine its ability to bind and neutralize the original (D614G) or Beta lineages. The first and second wave participants were hospitalized patients, who were closely matched in age with a median of 52 years (range 27-72) and 55 years (range 24-73), respectively. Samples were collected a median of 10 days (range 7-33) and 13 days (range 2-29) after a positive SARS-CoV-2 PCR test (Table S1). Although wave 1 viral sequences were not obtained, these samples were collected several months prior to the emergence of Beta in South Africa and were therefore assumed to have been infections caused by the original D614G lineage (Figure 1A). Wave 2 samples were collected when Beta accounted for >90% of infections in the region, with sequences from nasal swabs of 8/21 patients confirmed as Beta (Table S1), as described previously ^24^.

**Figure 1:**
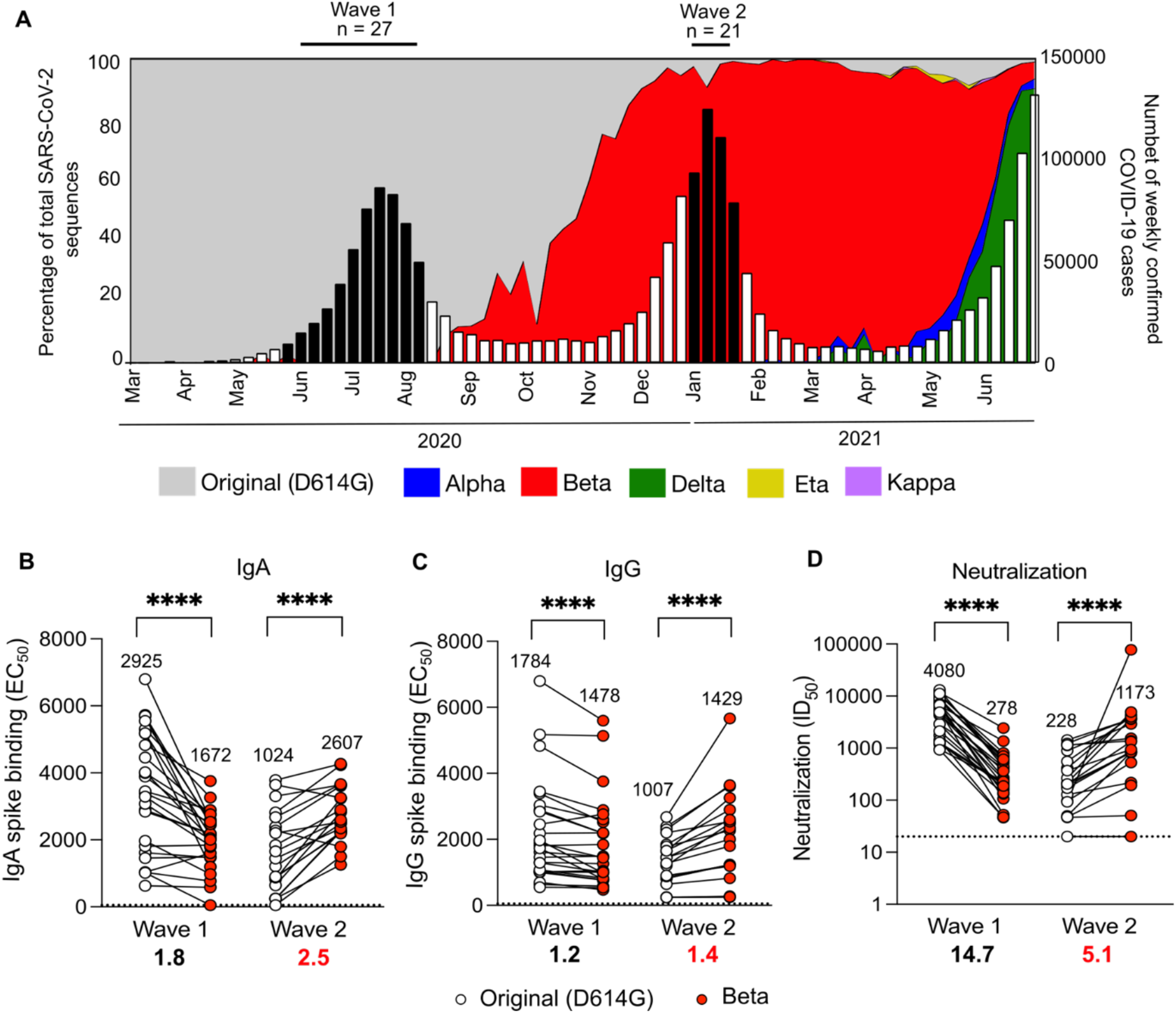
Binding and neutralization of plasma from hospitalized SARS-CoV-2 convalescent individuals sampled in waves driven by distinct viral lineages. (**A**) The number of SARS-CoV-2 cases in South Africa per epidemiological week from March 2020-July 2021 (right y axis) is represented by bars, with black bars indicating the period that the wave 1 samples (n=27) and wave 2 (n = 23) samples were obtained. The percentage of total SARS-CoV-2 sequences over time (left y axis) is shown as a line plot where the proportions of the original D614G, Alpha, Beta, Gamma, Delta, Eta and Kappa lineages are shown. (**B**) IgA and (**C**) IgG binding levels by ELISA of wave 1 or wave 2 samples against the original (D614G) (white) or Beta (red) spike. (**D**) Neutralization of original (D614G) or Beta pseudoviruses by wave 1 and 2 plasma. Limits of detection are shown with dotted lines, geometric mean titers (GMT) are shown and fold change decrease in black and fold change increase in red below the graph. Statistical significance between variants was calculated using Wilcoxon paired T test where **p<0.01; ****p<0.0001.

Comparison of the binding of IgA and IgG from Wave 1 plasma to the original (D614G) or Beta spikes showed a significant decrease in binding to Beta (Figure 1B and C). In contrast, wave 2 plasma from Beta infections showed a significant increase in both IgA and IgG binding to Beta. However, the differences in geometric mean titer against the original and Beta spike for wave 1 and 2 plasma were less than 2-fold for both IgG and IgA. In contrast, neutralization titers in plasma from wave 1 decreased 14.7 fold against Beta, while wave 2 titers of Beta were 5.1 fold higher than those of D614G (Figure 1C). This is in line with our previous studies using samples from the same cohorts ^6,24^. This difference in reduction of binding and neutralizing antibodies confirms the ability of convalescent plasma to target epitopes beyond the neutralizing epitopes mutated in VOCs.

### Cross-reactivity of Fc effector function is differentially impacted by the sequence of the infecting spike

We next measured whether Fc effector functions elicited in response to the original D614G variant or the Beta variant were equivalent in terms of magnitude and cross-reactivity. We measured spike-specific Fc responses using either spike protein coated on to fluorescent beads (for ADCP and ADCD) or spike expressed on the surface of target cells (ADCC and ADCT). In order to validate these assays, we tested the Fc effector function of monoclonal antibodies previously characterized for cell surface spike binding to D614G and Beta ^25^. A23-58.1, B1-182.1, A19-46.1 and A19-61.1 have previously been shown to bind D614G and Beta to similar levels, and therefore showed similar ADCP, ADCC, ADCD and ADCT activity against both D614G and Beta (Figure S1). Similarly, Class 4 mAb CR3022, which binds to regions of the RBD that exclude sites mutated in Beta ^26^, showed similar Fc effector function for both variants indicating that these assays were comparable across variants. For Class 2 mAbs BD23, LY-CoV555 and P2B-2F6, which are unable to bind or neutralize Beta ^6,27^, Fc effector function against Beta was similarly reduced.

Plasma from wave 1 participants all showed a significantly decreased ability to mediate ADCP, ADCC, ADCD and ADCT of Beta, compared to the original D614G spike, though all retained some activity against Beta (Figure 2A). However, not all Fc effector functions were affected to the same extent. ADCP, ADCC and ADCT showed 1.5, 1.4 and 1.4 fold decreases, similar to the 1.2 fold decrease observed for binding antibodies, however ADCD showed a 2.4 fold drop (Figure 2A and B). This suggests the possibility of epitope-specific functional responses. In contrast, wave 2 plasma from Beta infected individuals mediated similar levels of Fc function across the variants tested, with the exception of ADCC (Figure 2A), indicating the elicitation of more cross-reactive responses by this VOC. While wave 2 plasma samples mediated ADCP, ADCD and ADCT to similar levels against D614G and Beta spikes, ADCC was significantly higher against Beta (Figure 2A) (median D614G ADCC = 138, Beta ADCC = 249, 1.8 fold difference). The other Fc functions tested showed fold differences close to 1 for wave 2 samples (0.8, 0.9 and 1.1 for ADCP, ADCD and ADCT respectively) (Figure 2A and 2B).

**Figure 2:**
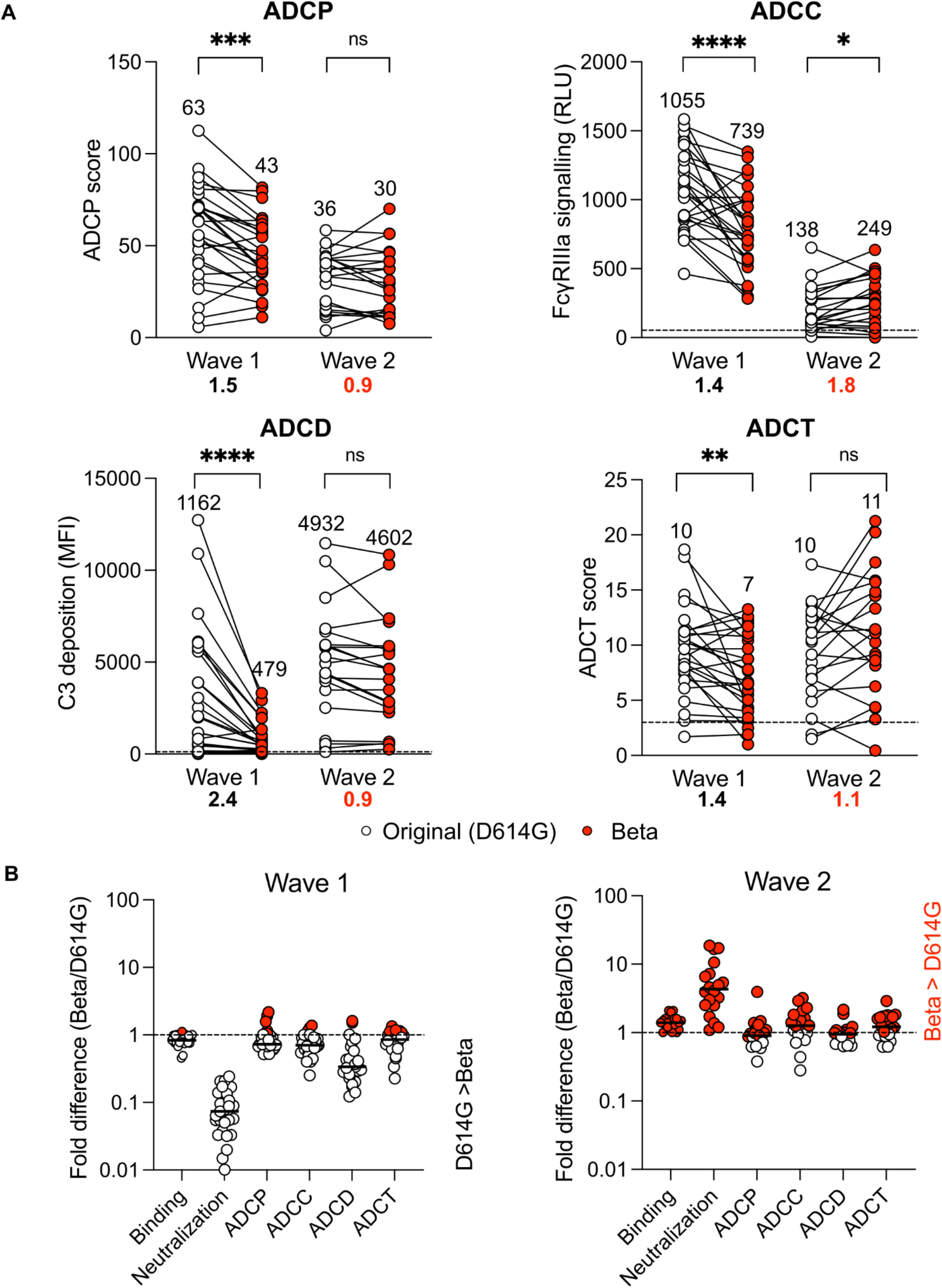
Fc effector function is largely preserved against Beta. (**A**) Fc effector functions of wave 1 and wave 2 plasma against either original (white) and Beta (red) spike protein or spike-expressing cells. Antibody-dependent cellular phagocytosis (ADCP) is represented as the percentage of monocytic cells that take up spike coated beads multiplied by their geometric mean fluorescence intensity (MFI). Antibody dependent cellular cytotoxicity (ADCC) shown as relative light units (RLU) signaling through Fcγ RIIIa expressing cells. Antibody dependent complement deposition (ADCD) is shown as the MFI of C3 deposition on spike coated beads and antibody dependent cellular trogocytosis (ADCT) represented as the relative proportion of biotinylated spike expressing cell membrane on CSFE+ monocytic cells. Dotted lines indicate the limit of detection and all samples are represented following the subtraction of background. Median values of all functions are shown above the graphs and fold change decrease in black and fold change increase in red below the graph. Statistical significance between variants is calculated using Wilcoxon paired T test where *p<0.05; **p<0.01; ***p<0.001; ****p<0.0001 and ns = non-significant. (**B**) Fold difference of functions against Beta relative to the original variant for wave 1 and 2 samples where the dotted line indicates no change between variants (red = Beta > D614G; white = D614G < Beta). The median of the fold changes is indicated by lines.

As Fc effector function is modulated through Fc receptor binding, we examined the ability of antibodies from both wave 1 and 2 to crosslink dimeric Fcγ receptors Fcγ RIIa or Fcγ RIIIa (which modulate ADCP and ADCC activity, respectively) and the original or Beta spike protein by ELISA. As expected, Spearman’s correlations exceeding 0.5 were noted between Fcγ RIIa binding and ADCP score, and between Fcγ RIIIa binding and ADCC against original D614G spike (Figure S2A and B). Similar to the functional readout, wave 1 samples showed significant decreases in their ability to mediate Beta specific Fcγ RIIa (Figure S2C) and Fcγ RIIIa cross-linking (Figure S2D), while no significant differences were noted for wave 2 samples (Figure S2E).

We considered the possibility that differences in Fc effector function simply reflected varying IgG levels between the waves, although the samples had been matched for age, severity and time since PCR test. Wave 1 and wave 2 samples showed no significant difference in IgG binding titers and ADCT activity to autologous spike (Figure 1C and 2A; Figure S2F). However, the wave 2 plasma showed significantly lower neutralization, ADCP and ADCC activity and enhanced ability to deposit complement protein compared to wave 1 plasma against autologous spike (Figure S2F). This illustrates that Fc effector VOC cross-reactivity is not as a result of binding titer. Overall, preserved Fc effector function, but substantial loss in neutralization against Beta in wave 1 samples (Figure 2B) suggests targeting by Fc effector function is distinct from that of neutralization.

### ADCC targets both the NTD and RBD regions of SARS-CoV-2 variants of concern

Although Fc effector function elicited by the original D614G virus was not completely abrogated in response to Beta, the significant decrease in activity suggests that NTD and RBD, which are mutated in Beta, are substantial targets. Given the significant decrease of ADCC observed against Beta by wave 1 plasma (Figure 2A) we sought to map these responses. Specifically, we determined the relative contribution to ADCC of antibodies targeting NTD and RBD by measuring Fcγ RIIIa signaling as a result of crosslinking to NTD or RBD proteins from the D614G and Beta variants.

First, we confirmed the ability of our assay to map these responses through the use of monoclonal antibodies. As for full spike (Figure S1), CR3022 ADCC against the RBD was unaffected by mutations in the Beta RBD (K417N, E484K and N501Y) while ADCC mediated by P2B-2F6 was abrogated (Figure 3A). 4A8, an NTD targeted mAb for which neutralization and binding is escaped by Beta (L18F, D80A, D215G and 242-244 del) ^6^ showed ADCC activity against the original variant but not Beta RBD (Figure 3B).

**Figure 3:**
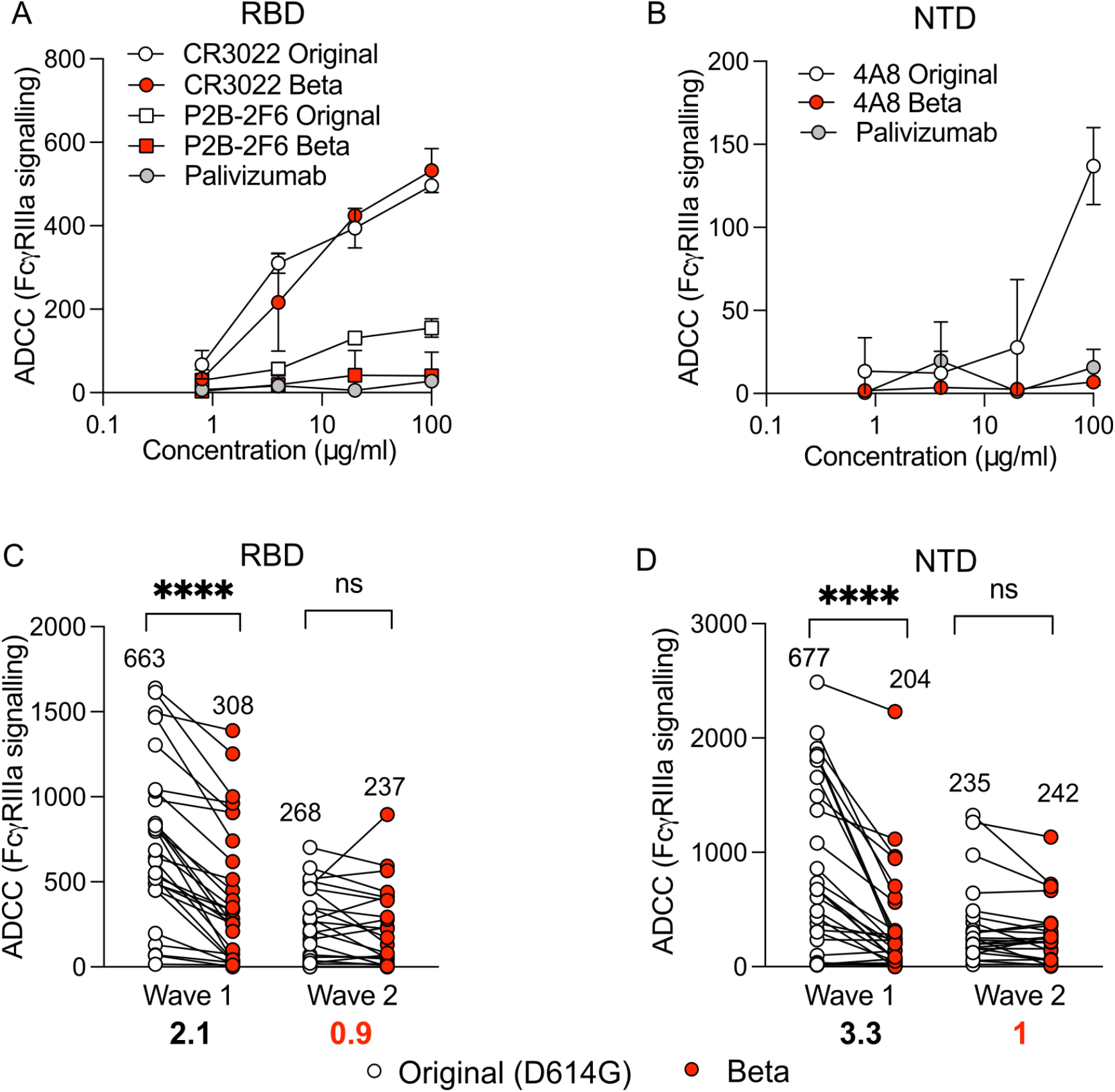
ADCC targets the Receptor binding domain (RBD) and N-Terminal Domain (NTD) and is partially escaped by Beta. (**A**) ADCC of monoclonal antibodies CR3022, P2B-2FB and Palivizumab shown as RLU of signaling through Fcγ RIIIa expressing cells and cross-linking of original (white) or Beta (red) (K417N, E484K and N501Y) RBD protein. (**B**) ADCC of monoclonal antibodies 4A8 and Palivizumab against original (white) or Beta (red; L18F, D80A, D215G, 242-244 del) NTD protein. Wave 1 and Wave 2 plasma against original (white) or Beta (red) (**C**) RBD or (**D**) NTD protein. All plots are representative of a minimum of two independent experiments. Median values of all functions are shown above the graphs with fold change decrease in black and fold change increase in red below the graph. Statistical significance between variants was calculated using Wilcoxon paired T test where ****p<0.0001 and ns = non-significant.

ADCC mediated by wave 1 plasma showed a 2.2 fold decrease against Beta RBD (Median 663 and 308 for original and Beta respectively) confirming that while K417, E484 and N501 sites are targeted, they do not account for the majority of ADCC activity against the RBD (Figure 3C). Beta-elicited ADCC did not show significant differences between the original or Beta RBD (Figure 3C), which may suggest broader tolerance of RBD mutations in wave 2 plasma, consistent with neutralization. Similarly, ADCC was detected against the original NTD protein with a 3 fold decrease (median 677 to 204) against Beta in Wave 1 plasma (Figure 3C). This may indicate that ADCC antibodies more frequently target NTD sites mutated in Beta (L18, D80, D215), or are less able to tolerate the conformation change of the NTD that may result from the 242-244 deletion. Conversely, as for RBD, ADCC elicited by Beta was not significantly different against the original or Beta NTD (Figure 3C). Overall these data show that NTD and RBD are targets of ADCC responses in convalescent plasma. However, mutations in these key regions that allow for complete neutralization escape in individuals infected with the original (D614G) variant, only slightly affect ADCC.

### Beta-elicited ADCC shows greater cross-reactivity against a panel of global variants of concern compared to wave 1 infection and Ad26.COV2.S vaccination

As Beta-elicited plasma showed enhanced cross-reactivity for the original variants, we assessed a larger panel of global VOCs using cells expressing spikes from the original D614G and Alpha, Beta, Gamma, Delta and SARS-1 as targets. Given the transient nature of the transfections, different variants were run head-to-head and normalized by CR3022 ADCC activity.

For wave 1 sera, despite high levels of ADCC against the autologous original variant (Median 1055), the capacity of antibodies to mediate ADCC against all heterologous spikes except for Alpha (median 841) was significantly reduced (Beta: 739; Gamma: 635 and Delta: 662) (Figure 4A). The most dramatic reduction in activity was observed against SARS1, which differed significantly from all other spikes (median: 228). In contrast, Wave 2 plasma showed similar ADCC levels across SARS-CoV-2 variants (Figure 4B) with Gamma-specific ADCC showing the highest level (median 284), followed by Beta, Delta, Alpha and the original variant (250,198,159 and 138 respectively). SARS-1 specific ADCC, while detectable, was significantly lower than ADCC directed against all SARS-CoV-2 variants as expected. These data indicate that Beta-elicited ADCC activity is cross-reactive with other VOCs, whereas ADCC in response to the original (D614G) variant was substantially less cross-reactive.

**Figure 4:**
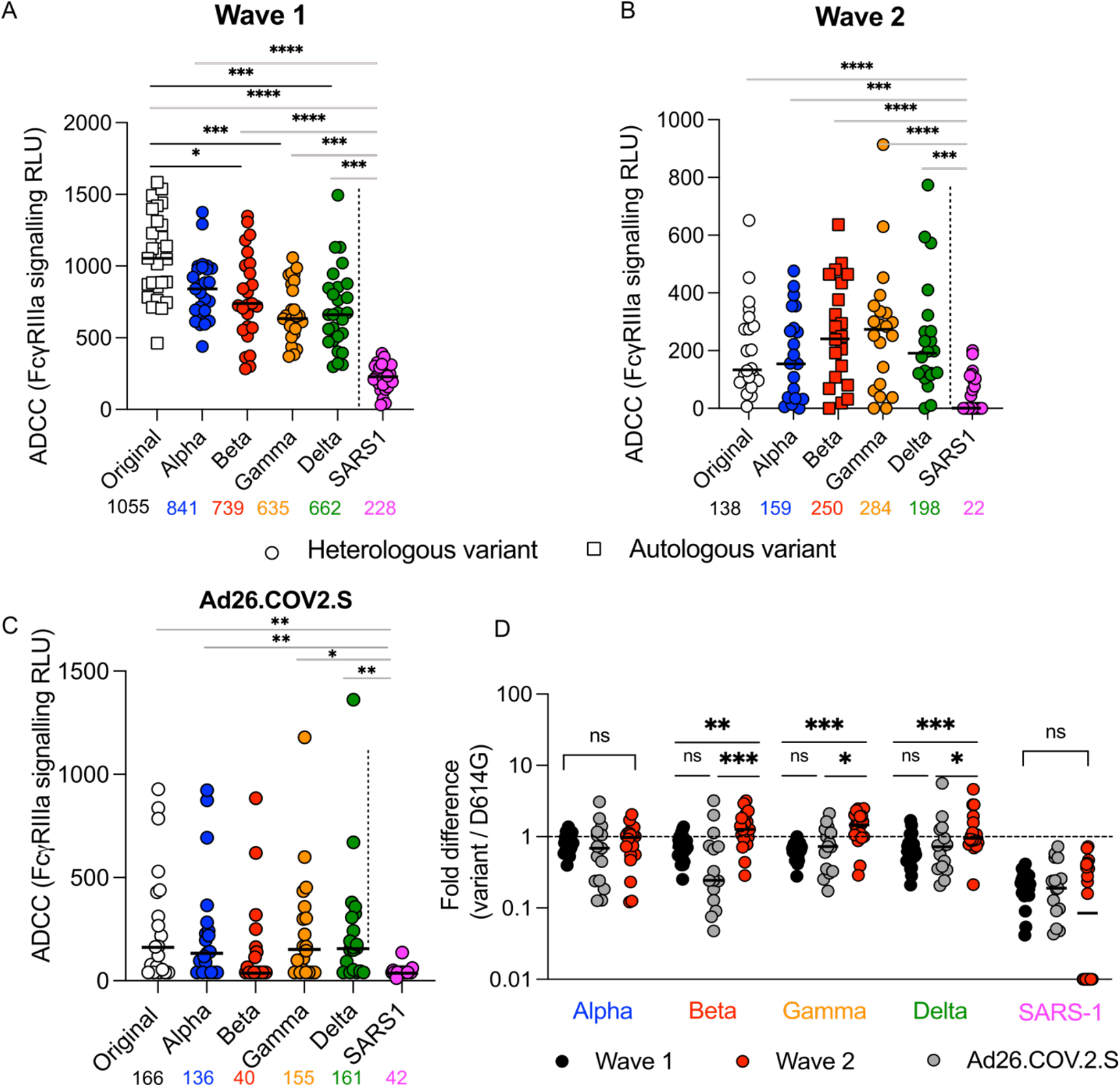
Beta and Ad26.COV.2.S-elicited ADCC responses are cross-reactive against variants of concern. ADCC of (**A**) wave 1, (**B**) wave 2 and (**C**) Ad26.COV.2.S vaccinee plasma (day 28) against the original, VOC and SARS-CoV-1. Bars indicate the median, represented below the graphs with squares indicating the ADCC against the autologous variant and circles indicating ADCC against the heterologous variants. Statistical significance was calculated using the Friedman test, with Dunn’s multiple test comparisons, where grey lines indicate significance between SARS-CoV-2 and SARS-CoV-1 and black lines between SARS-CoV-2 variants. (**D**) Fold difference represented as ADCC activity against the variant for wave 1, wave 2 or vaccine plasma relative to the original. The dotted line represents no fold difference while lines indicate the median. Statistical differences between waves and vaccine responses were calculated using the Kruskal-Wallis test with Dunn’s multiple test comparisons. *p<0.05; ***p<0.001; ****p<0.0001 and ns = non-significant.

In order to contextualize the ADCC VOC cross-reactivity seen in infection, we compared this to Janssen/Johnson and Johnson Ad26.COV.2.S vaccination that was rolled out in South Africa shortly after the second wave. ADCC was measured 28 days post vaccination in 19 individuals previously confirmed to be SARS-CoV-2 unexposed ^9^. Beta was the most ADCC resistant of the VOCs tested, showing a 3-fold decrease compared to the original variant (median of 40 compared to 166), whereas ADCC against all other variants showed similar or less than 2-fold decreases (Figure 4C) (fold change relative to the original: Alpha 1.2; Gamma 1.1; Delta 1). While responses against SARS-1 were significantly lower than most VOCs, they were detectable in some participants.

Vaccine and wave 1 ADCC against VOCs were similar, indicating comparable levels of cross reactivity (Figure 4D). Strikingly though, vaccine-elicited plasma showed decreased capacity to perform ADCC against Beta compared to wave 1 plasma (Figure 4D). For Beta, Gamma and Delta, the fold differences for wave 2 sera were consistently >1, and significantly different from wave 1 and Ad26.COV2.S, while Alpha and SARS-1 showed no difference (Figure 4D). This indicates that overall, Beta-elicited ADCC showed greater cross-reactivity to global VOCs compared to original elicited ADCC by either vaccination or infection.

## Discussion

The emergence of variants that can escape neutralizing responses has compromised vaccine protection from infection, but in many cases not from severe disease. This has highlighted the potential role of other immune functions, including Fc effector functions in mitigation of disease severity. Previous studies have shown sustained activity against VOCs for both convalescent donors and vaccinees ^7,10,23^. Here, we confirm that while there is overall preservation of Fc effector function against VOCs in infection and vaccination, the magnitude of Fc effector activity against Beta is reduced. Furthermore, escape varies by function, suggesting that antibodies mediating different Fc functions may have distinct epitopes. We also assessed Fc function in convalescent individuals who were infected with a VOC. We show that, compared to either the original variant or Ad26.COV2.S vaccination, infection with Beta imprints significantly improved Fc cross-reactivity against global VOCs, and that these antibodies target epitopes distinct from those of neutralizing antibodies.

Several studies now support the notion that unlike neutralization, Fc effector function is more resilient in the face of VOCs ^7,23^. In this study we showed preservation of Fc effector function against VOCs both in individuals previously infected with the original D614G variant but also in individuals vaccinated with Ad26.COV2.S. This is supported by a recent study measuring NK activation, ADCP, ADCD and ADCC in Ad26.COV2.S vaccinees showing differences of under 2-fold between original D614G and Beta^2^. Further, this is translatable for NK activation and ADCP across other vaccine modalities including mRNA-1273 and BNT162b2^7,23^.

While Fc effector functions against Beta were only slightly lower, loss of activity was not equivalent for all functions, with ADCD being most affected. RBD is a major target for complement binding in both vaccinated and convalescent individuals ^28^, which may explain why this function in particular has greater loss against Beta, which has key immune evasion mutations in the RBD. In contrast to ADCD, ADCC activity against Beta was less reduced.. Our epitope mapping data indicates that Beta RBD and NTD are not the predominant targets of this function. Therefore preservation of ADCC may be the result of antibodies targeting epitopes or sites beyond those commonly mutated in VOCs. Differential epitope targeting has also been suggested for other functions, with RBD depletion greatly decreasing ADNP activity in contrast to ADCP, which remained unaffected in convalescent plasma ^23^. In addition to antibody targeting that varied by function, we also show that individuals infected by different variants have unique Fc effector profiles despite similar levels of binding antibodies. These data suggest differential targeting of individual Fc functions in response to VOC. Further delineation of the targets of these responses may allow manipulation of future immunogens to specifically improve Fc effector functions.

We also show subtle differences in the ability of antibodies elicited by either the original D614G or the Ad26.COV2.S vaccine to perform ADCC against Beta. This was despite the fact that the sequence of immunodominant regions of the eliciting immunogens were the same, with only the single D614G mutation differing between them. Similar findings have been reported for ADCP, where RBD is targeted to varying levels in different vaccine modalities and convalescent plasma ^23^. This suggests that beyond sequence, nuanced differences in antigen stability and presentation affect functional responses to vaccination or infection.

We have previously shown that Beta infection imprints a cross-reactive neutralizing response ^3,24^. Here we show that this also extends to Fc effector function, suggesting that features intrinsic to each spike shape the overall antibody repertoire. However, the pattern of breadth between neutralization and Fc effector function is not necessarily the same for a given spike. For neutralization, comparison of convalescent sera from Beta and Delta infections have suggested that these VOCs are serologically distant, with Beta-elicited plasma showing diminished ability to neutralize Delta ^29,30^. However, for ADCC, we find that cross reactivity is enhanced against Delta by Beta-elicited plasma. This further points to the divergent impact that sequence of immunogen may have on neutralization and Fc effector function.

Collectively these data show the persistence of Fc effector function against emerging VOCs in the face of waning neutralization responses. It further suggests that the spike sequence of the priming immunogen is likely to determine unique Fc effector function profiles, allowing for their potential modulation in future vaccine design. This however should be considered in the context of neutralization for which the choice of immunogen sequence is far more constrained and alongside which Fc effector function is likely to play a supporting role in protection. While current vaccines provide sufficient protection against severe disease, vaccination strategies may be improved by a spike immunogen associated with a more balanced and broad response.

## Data Availability

All data produced in the present study are available upon reasonable request to the authors

## Acknowledgements

We thank Dr Bronwen Lambson, Donald Mhlanga and Brent Oosthuysen for generating viral variants of concern. We thank Mashudu Madzivhandila and Thandeka Moyo-Gwete for generating the initial wave 2 neutralization and binding data. We thank Amkele Ngomti, Richard Baguma and Roanne Keeton for assistance with sample processing. We thank Mieke van der Mescht, Zelda van der Walt, Talita de Villiers, Fareed Abdullah, Paul Rheeder, Albertus Malan, and Wesley van Hougenhouck-Tulleken for contributing to patient management, sample collection and processing, and data management for the Pretoria COVID-19 study. We thank the study participants at Groote Schuur Hospital and Steve Biko Academic Hospital and Elloise du Toit for assistance with clinical data and Nigel Garrett and Ameena Goga and the Sisonke vaccination team. The parental soluble spike was provided by Jason McLellan (University of Texas). The parental pseudovirus plasmids were kindly provided by Drs Elise Landais and Devin Sok (IAVI). W.A.B. is supported by the EDCTP2 programme of the European Union’s Horizon 2020 programme (TMA2016SF-1535-CaTCH-22) and Wellcome Centre for Infectious Diseases Research in Africa (CIDRI-Africa), which is supported by core funding from the Wellcome Trust (203135/Z/16/Z). N.A.B.N acknowledges funding from the SA-MRC, MRC UK, NRF and the Lily and Ernst Hausmann Trust. PLM is supported by the South African Research Chairs Initiative of the Department of Science and Innovation and National Research Foundation of South Africa, the SA Medical Research Council SHIP program, the Centre for the AIDS Program of Research (CAPRISA). SIR is a L’Oreal/UNESCO Women in Science South Africa Young Talents awardee. Related research by the authors is conducted as part of the DST-NRF Centre of Excellence in HIV Prevention, which is supported by the Department of Science and Technology and the National Research Foundation.

## Author contributions

S.I.R designed the study, performed experiments, analyzed the data and wrote the manuscript. N.P.M and B.M.M performed Fc experiments and analyzed data. H.K and M.M performed neutralization assays. F.A and Z.M performed ELISA assays. L.G.B and G.G.G established the Sisonke Ad26.COV2.S trial. V.U, T.R. and M.B established the Pretoria COVID-19 study which provided participant samples from the Steve Biko Academic Hospital. M.M., S.S. and N.W. collected samples for and N.A.B.N and W.A.B established the Groote Schuur Hospital cohort. N.J.S. and J.M. provided monoclonal antibodies and PLM wrote the manuscript.

## Materials and Methods

### Human Subjects

First wave plasma samples (n = 27) were collected from participants enrolled to the Pretoria COVID-19 study cohort. Participants were admitted to Steve Biko Academic Hospital (Pretoria, South Africa) with moderate to severe (WHO scale 4-6) PCR confirmed SARS-CoV-2 infection between May and September 2020 (Figure 1A; Table S1). Ethics approval was received from the University of Pretoria, Human Research Ethics Committee (Medical) (247/2020). Second wave plasma samples were obtained from hospitalized COVID-19 patients (n=21) with moderate disease (WHO scale 4-5) admitted to Groote Schuur Hospital cohort, Cape Town from December 2020 – January 2021 (Figure 1A; Table S1). This study received ethics approval from the Human Research Ethics Committee of the Faculty of Health Sciences, University of Cape Town (R021/2020). All patients had polymerase chain reaction (PCR) confirmed SARS-CoV-2 infection a median of 7 days (IQR 3-11) before blood collection. In both cohorts, clinical folders were consulted for participants and none showed evidence of prior symptomatic COVID-19 disease or previous positive PCR test results. For the Ad26.COV.2 vaccine samples (n = 19), health care workers were recruited between July 2020 and January 2021 from Groote Schuur Hospital (Cape Town, Western Cape, South Africa) and vaccinated with single dose Johnson and Johnson Ad26.COV2.S in the Sisonke Phase 3b trial between 12 February and 26 March 2021. The lack of prior infection in these individuals was confirmed by Nucleocapsid ELISA as previously described ^9^. The study was approved by the University of Cape Town Human Research Ethics Committee (HREC 190/2020 and 209/2020) and the University of the Witwatersrand Human Research Ethics Committee (Medical) (no M210429). Written informed consent was obtained from all participants.

### Cell lines

Human embryo kidney HEK293T cells were cultured at 37°C, 5% CO_2_, in DMEM containing 10% heat-inactivated fetal bovine serum (Gibco BRL Life Technologies) and supplemented with 50 μg/ml gentamicin (Sigma). Cells were disrupted at confluence with 0.25% trypsin in 1 mM EDTA (Sigma) every 48–72 hours. HEK293T/ACE2.MF cells were maintained in the same way as HEK293T cells but were supplemented with 3 μg/ml puromycin for selection of stably transduced cells. HEK293F suspension cells were cultured in 293 Freestyle media (Gibco BRL Life Technologies) and cultured in a shaking incubator at 37°C, 5% CO2, 70% humidity at 125rpm maintained between 0.2 and 0.5 million cells/ml. Jurkat-Lucia™ NFAT-CD16 cells were maintained in IMDM media with 10% heat-inactivated fetal bovine serum (Gibco, Gaithersburg, MD), 1% Penicillin Streptomycin (Gibco, Gaithersburg, MD) and 10 μg/ml of Blasticidin and 100 μg/ml of Zeocin was added to the growth medium every other passage. THP-1 cells were used for both the ADCP and ADCT assays and obtained from the AIDS Reagent Program, Division of AIDS, NIAID, NIH contributed by Dr. Li Wu and Vineet N. KewalRamani. Cells were cultured at 37°C, 5% CO2 in RPMI containing 10% heat-inactivated fetal bovine serum (Gibco, Gaithersburg, MD) with 1% Penicillin Streptomycin (Gibco, Gaithersburg, MD) and 2-mercaptoethanol to a final concentration of 0.05 mM and not allowed to exceed 4 × 10^5^ cells/ml to prevent differentiation.

### SARS-CoV-2 spike genome sequencing

For wave 2 samples, sequencing of the spike was performed as previously described ^24^ using swabs obtained from 28 randomly collected Groote Schuur Hospital patients of which eight were included in this study. RNA sequencing was performed as previously published ^1^. Briefly, extracted RNA was used to synthesize cDNA using the Superscript IV First Strand synthesis system (Life Technologies, Carlsbad, CA) and random hexamer primers. SARS-CoV-2 whole genome amplification was performed by multiplex PCR using primers designed on Primal Scheme (http://primal.zibraproject.org/) to generate 400 bp amplicons with a 70 bp overlap covering the SARS-CoV-2 genome. Phylogenetic clade classification of the genomes in this study consisted of analyzing them against a global reference dataset using a custom pipeline based on a local version of NextStrain (https://github.com/nextstrain/ncov) ^31^.

### SARS-CoV-2 antigens

For ELISA, ADCP and ADCD assays, SARS-CoV-2 original and Beta variant full spike (L18F, D80A, D215G, K417N, E484K, N501Y, D614G, A701V, 242-244 del), RBD original and Beta (K417N, E484K and N501Y, D614G) and NTD original and Beta (L18F, D80A, D215G and 242-244 del) proteins were expressed in Human Embryonic Kidney (HEK) 293F suspension cells by transfecting the cells with the respective expression plasmid. After incubating for six days at 37 °C, 70% humidity and 10% CO_2_, proteins were first purified using a nickel resin followed by size-exclusion chromatography. Relevant fractions were collected and frozen at -80 °C until use.

### SARS-CoV-2 Spike Enzyme-linked immunosorbent assay (ELISA)

Two μg/ml of spike protein (Original or Beta) was used to coat 96-well, high-binding plates and incubated overnight at 4 °C. The plates were incubated in a blocking buffer consisting of 5% skimmed milk powder, 0.05% Tween 20, 1x PBS. Plasma samples were diluted to 1:100 starting dilution in a blocking buffer and added to the plates. IgG or IgA secondary antibody was diluted to 1:3000 or 1:1000 respectively in blocking buffer and added to the plates followed by TMB substrate (Thermofisher Scientific). Upon stopping the reaction with 1 M H_2_SO_4_, absorbance was measured at a 450nm wavelength. In all instances, mAbs CR3022 and BD23 were used as positive controls and Palivizumab was used as a negative control.

### Spike plasmid and Lentiviral Pseudovirus Production

The SARS-CoV-2 Wuhan-1 spike, cloned into pCDNA3.1 was mutated using the QuikChange Lightning Site-Directed Mutagenesis kit (Agilent Technologies) and NEBuilder HiFi DNA Assembly Master Mix (NEB) to include D614G (original) or lineage defining mutations for Beta (L18F, D80A, D215G, 242-244del, K417N, E484K, N501Y, D614G and A701V), Gamma (L18F, T20N, P26S, D138Y, R190S, K417T, E484K, N501Y, D614G, H655Y, T1027I, V1176F) or Delta (T19R, 156-157del, R158G, L452R, T478K, D614G, P681R and D950N). SARS-1 spike was also cloned into pcDNA.

Pseudotyped lentiviruses were prepared by co-transfecting HEK293T cell line with either the SARS-CoV-2 ancestral variant spike (D614G) or the Beta spike plasmids in conjunction with a firefly luciferase encoding lentivirus backbone plasmid as previously described ^24^. Briefly, pseudoviruses were produced by co-transfection in 293T/17 cells with a lentiviral backbone (HIV-1 pNL4.luc encoding the firefly luciferase gene) and either of the SARS-CoV-2 spike plasmids with PEIMAX (Polysciences). Culture supernatants were clarified of cells by a 0.45-μM filter and stored at ™70 °C. Other pcDNA plasmids were used for the ADCC assay.

### Pseudovirus neutralization assay

For the neutralization assay, plasma samples were heat-inactivated and clarified by centrifugation. Heat-inactivated plasma samples from vaccine recipients were incubated with the SARS-CoV-2 pseudotyped virus for 1 hour at 37°C, 5% CO2. Subsequently, 1×10^4^ HEK293T cells engineered to over-express ACE-2 (293T/ACE2.MF)(kindly provided by M. Farzan (Scripps Research)) were added and incubated at 37°C, 5% CO_2_ for 72 hours upon which the luminescence of the luciferase gene was measured. Titers were calculated as the reciprocal plasma dilution (ID50) causing 50% reduction of relative light units. CB6 and CA1 was used as a positive control.

### Antibody-dependent cellular phagocytosis (ADCP) assay

SARS-CoV-2 original or Beta spike was biotinylated using EZ link Sulfo-NHS-LC-Biotin kit (ThermoFisher) and coated on to fluorescent neutravidin beads as previously described ^32^. Briefly, beads were incubated for two hours with monoclonal antibodies at a starting concentration of 2 μg/ml and titrated five-fold or plasma at a single 1 in 100 dilution. Opsonized beads were incubated with the monocytic THP-1 cell line overnight, fixed and interrogated on the FACSAria II. Phagocytosis score was calculated as the percentage of THP-1 cells that engulfed fluorescent beads multiplied by the geometric mean fluorescence intensity of the population less the no antibody control. For this and all subsequent Fc effector assays, pooled plasma from 5 PCR-confirmed SARS-CoV-2 infected individuals and CR3022 were used as positive controls and plasma from 5 pre-pandemic healthy controls and Palivizumab were used as negative controls. ADCP scores for original and Beta spikes were normalised to each other and between runs using CR3022.

### Antibody-dependent cellular cytotoxicity (ADCC) assay

The ability of plasma antibodies to cross-link and signal through FcγRIIIa (CD16) and spike expressing cells or SARS-CoV-2 protein was measured as a proxy for ADCC. For spike assays, HEK293T cells were transfected with 5μg of SARS-CoV-2 original variant spike (D614G), Beta, Gamma, Delta or SARS-1 spike plasmids using PEI-MAX 40,000 (Polysciences) and incubated for 2 days at 37°C. Expression of spike was confirmed by differential binding of CR3022 and P2B-2F6 and their detection by anti-IgG APC staining measured by flow cytometry. For original or Beta NTD or RBD assays protein was coated at 1 μg/ml on a high binding ELISA 96-well plate and incubated at 4°C overnight. Plates were then washed with PBS and blocked at room temperature for 1 hr with PBS + 2.5% BSA. Subsequently, protein or 1×10^5^ spike transfected cells per well were incubated with heat inactivated plasma (1:100 final dilution) or monoclonal antibodies (final concentration of 100 μg/ml) in RPMI 1640 media supplemented with 10% FBS 1% Pen/Strep (Gibco, Gaithersburg, MD) for 1 hour at 37°C. Jurkat-Lucia™ NFAT-CD16 cells (Invivogen) (2×10^5^ cells/well and 1×10^5^ cells/well for spike and other protein respectively) were added and incubated for 24 hours at 37°C, 5% CO_2_. Twenty μl of supernatant was then transferred to a white 96-well plate with 50 μl of reconstituted QUANTI-Luc secreted luciferase and read immediately on a Victor 3 luminometer with 1s integration time. Relative light units (RLU) of a no antibody control was subtracted as background. Palivizumab was used as a negative control, while CR3022 was used as a positive control, and P2B-2F6 to differentiate the Beta from the D614G variant. To induce the transgene 1x cell stimulation cocktail (Thermofisher Scientific, Oslo, Norway) and 2 μg/ml ionomycin in R10 was added as a positive control to confirm sufficient expression of the Fc receptor. CR3022 (for spike and RBD) or 4A8 (NTD) were used as positive controls and Palivizumab were used as negative controls. RLUs for original and Beta spikes were normalised to each other and between runs using CR3022. A cut off of 40 was determined by screening of 40 SARS-CoV-2 naive and unvaccinated individuals.

### Antibody-dependent complement deposition (ADCD) assay

ADCD was measured as previously described ^33^. Biotinylated spike protein was coated 1:1 onto red fluorescent 1 μM neutravidin beads (Molecular Probes Inc.) for 2 hours at 37°C. These were incubated with a single 1:10 plasma sample dilution or 5-fold titration of mAb at a starting concentration of 100 μg/ml for 2 hours and guinea pig complement diluted 1 in 50 with gelatin/veronal buffer for 15 minutes at 37°C. Beads were washed in PBS and stained with anti-guinea pig C3b-FITC, fixed and interrogated on a FACS Aria II. Complement deposition score was calculated as the percentage of C3b-FITC positive beads multiplied by the geometric mean fluorescent intensity of FITC in this population less the no antibody or heat inactivated controls. ADCD scores for original and Beta spikes were normalised to each other and between runs using CR3022.

### Antibody-dependent cellular trogocytosis (ADCT) assay

ADCT was performed as described in and modified from a previously described study^34^. HEK293T cells transfected with a SARS-CoV-2 spike pcDNA vector as above were surface biotinylated with EZ-Link Sulfo-NHS-LC-Biotin as recommended by the manufacturer. Fifty-thousand cells per well were incubated with 5-fold titration of mAb starting at 25 μg/ml or single 1 in 100 dilution for 30 minutes. Following a RPMI media wash, these were then incubated with carboxyfluorescein succinimidyl ester (CFSE) stained THP-1 cells (5 ×10^4^ cells per well) for 1 hour and washed with 15mM EDTA/PBS followed by PBS. Cells were then stained for biotin using Streptavidin-PE and read on a FACSAria II. Trogocytosis score was determined as the proportion of CFSE positive THP-1 cells also positive for streptavidin-PE less the no antibody control.

### Dimeric Fc Gamma Receptor Binding ELISAs

High-binding 96 well ELISA plates were coated with 1 ug/ml spike protein in PBS overnight at 4°C. Three wells on each plate were directly coated with 5 ug/ml IgG, isolated from healthy donors, and signals from these wells were used to normalize the Fc receptor activity of the plasma samples. Plates were washed with PBS and blocked with PBS/1 mM EDTA/1% BSA for 1 hour at 37°C. Plates were then washed and incubated with 1:10 diluted plasma for 1 hour at 37°C and then with 0.2 ug/ml or 0.1 ug/ml of biotinylated FcγRIIa or FcγRIIIa dimer respectively (constructs kindly provided by Prof. Mark Hogarth from the Burnet Institute, Australia) for 1 hour at 37°C ^35^. Subsequently, a 1:10,000 dilution of Pierce high-sensitivity streptavidin-horseradish peroxidase (Thermo Scientific) was added for a final incubation for 1 hour at 37°C. Lastly, TMB substrate (Sigma-Aldrich) was added, and color development was stopped with 1 M H_2_SO_4_ and absorbance read at 450 nm.

### Statistical analysis

Analyses were performed in Prism (v9; GraphPad Software Inc, San Diego, CA, USA). Non-parametric tests were used for all comparisons. The Mann-Whitney and Wilcoxon tests were used for unmatched and paired samples, respectively. The Friedman test with Dunns correction for multiple comparisons was used for matched comparisons across variants. All correlations reported are non-parametric Spearman’s correlations. *P* values less than 0.05 were considered to be statistically significant.

## Supplementary data

**Figure S1: Fc effector function of monoclonal antibodies against original and Beta variants**

Titrations of ADCP, ADCC, ADCT and ADCD of SARS-CoV-2 binding antibodies and RSV specific Palivizumab against the original (D614G,white) and Beta (red) spike are shown as well as the area under the curve for each function indicated by bars. In all cases error bars represent the standard deviation of a minimum of 2 independent experiments.

**Figure S2: Fc**γ **receptor binding correlates with functional activity against SARS-CoV-2 variants**

Spearman’s correlations of (A) FcγRIIa dimer binding by ELISA and ADCP activity and (B) FcγRIIIa dimer binding by ELISA and ADCC activity of wave 1 and 2 plasma against spike where ***p<0.001, ****p <0.0001. (C) FcγRIIa and (D) FcγRIIIa dimer binding of wave 1 and wave 2 plasma against either original (white) and Beta (red) expressed spike protein. Statistical significance between variants calculated using Wilcoxon paired T test where ****p<0.0001 and ns = non-significant. (E) Fold difference of functions against Beta relative to the original variant for wave 1 and wave samples where the dotted line indicates no change between variants. The median of the fold changes are indicated by lines, with white dots. Indicating cases where D614G < Beta function and red dots where Beta > D614G. (F) Comparison of functional activity of wave 1 and wave 2 plasma against their respective autologous variants (original and Beta respectively). Neutralization as represented as IC50 titer, binding as EC50, ADCP as ADCP score, ADCC as FcγRIIIa signalling RLU, ADCD as MFI od C3 deposition and ADCT as ADCT score. Statistical differences between wave 1 and 2 were calculated by Mann Whitney t test where *p<0.05; ***p<0.001; ****p<0.0001 and ns = non-significant and bars indicate the median activity.

**Table S1: Demographic and clinical description of the wave 1 and wave 2 cohorts**

## Notes

### Competing Interest Statement

The authors have declared no competing interest.

### Funding Statement

W.A.B. is supported by the EDCTP2 programme of the European Unions Horizon 2020 programme (TMA2016SF-1535-CaTCH-22) and Wellcome Centre for Infectious Diseases Research in Africa (CIDRI-Africa), which is supported by core funding from the Wellcome Trust (203135/Z/16/Z). N.A.B.N acknowledges funding from the SA-MRC, MRC UK, NRF and the Lily and Ernst Hausmann Trust. PLM is supported by the South African Research Chairs Initiative of the Department of Science and Innovation and National Research Foundation of South Africa, the SA Medical Research Council SHIP program, the Centre for the AIDS Program of Research (CAPRISA). SIR is a LOreal/UNESCO Women in Science South Africa Young Talents awardee. Related research by the authors is conducted as part of the DST-NRF Centre of Excellence in HIV Prevention, which is supported by the Department of Science and Technology and the National Research Foundation.

### Author Declarations

Ethics approval was received from the University of Pretoria, Human Research Ethics Committee (Medical) (247/2020), the Human Research Ethics Committee of the Faculty of Health Sciences, University of Cape Town (R021/2020). The study was also approved by the University of Cape Town Human Research Ethics Committee (HREC 190/2020 and 209/2020) and the University of the Witwatersrand Human Research Ethics Committee (Medical) (no M210429).

