## Supplementary material for "A SARS-CoV-2 variant of concern triggers Fc effector function with increased cross-reactivity"

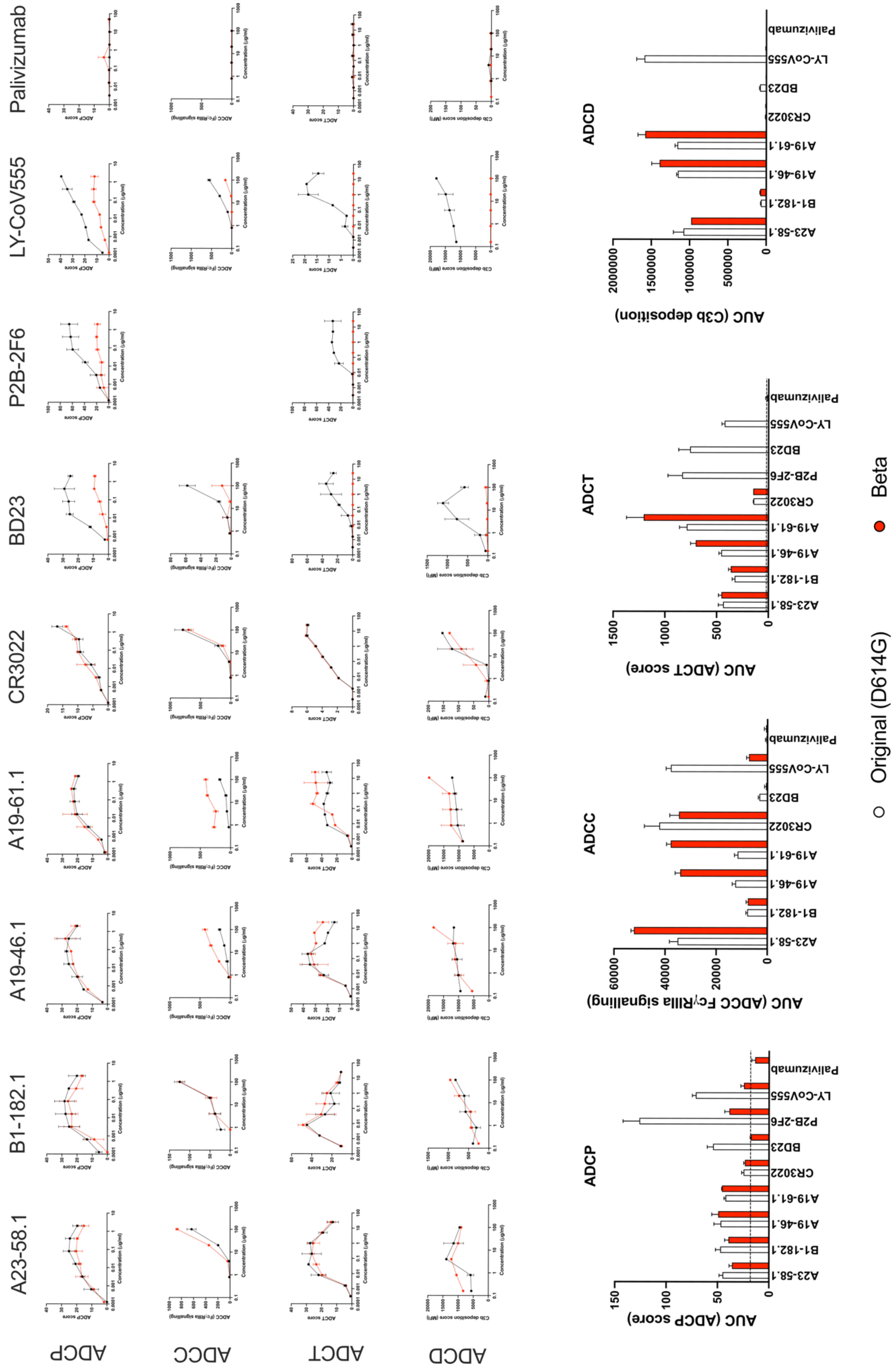


**Figure S1: Fc effector function of monoclonal antibodies against original and Beta variants**

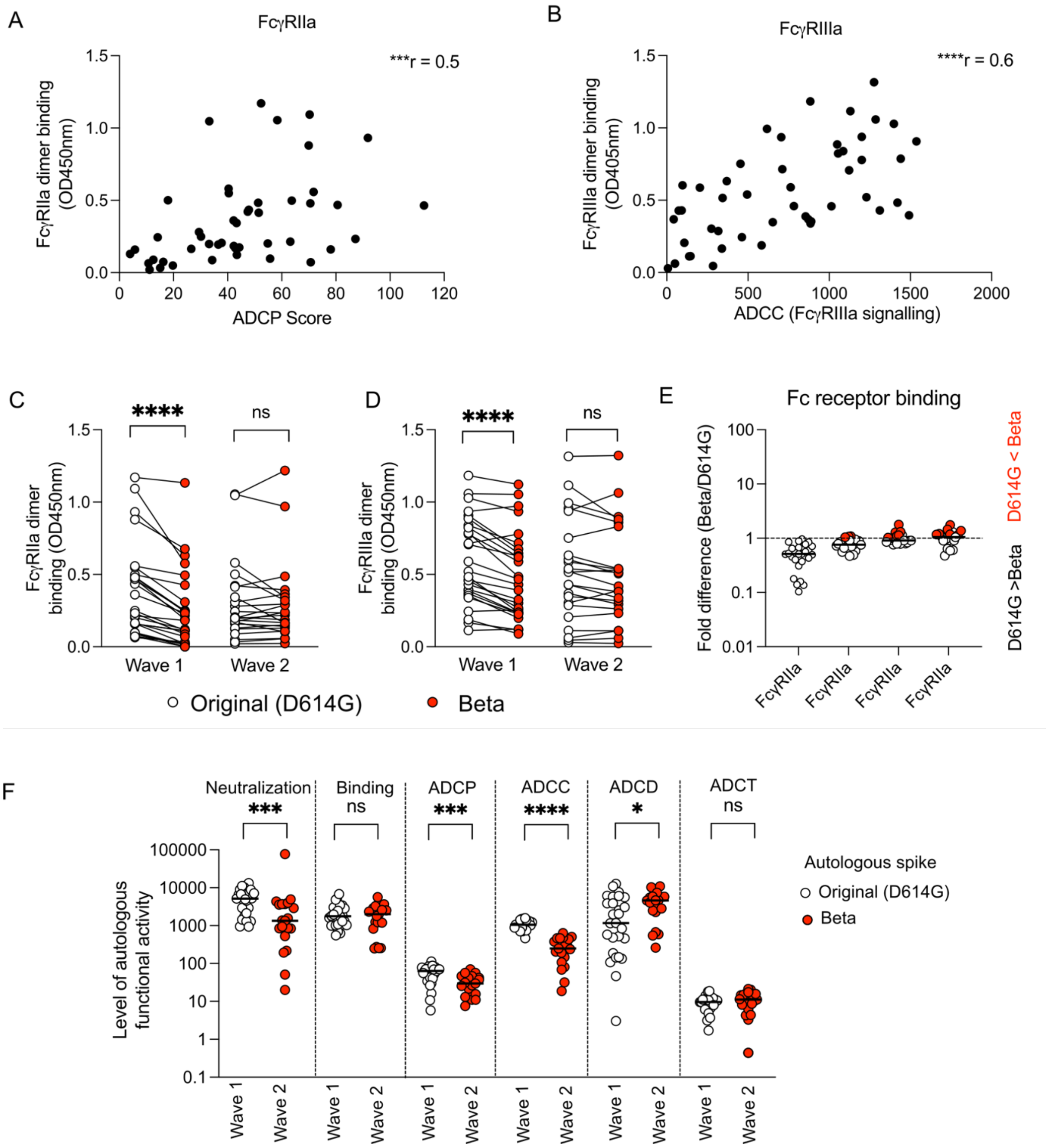


**Figure S2: Fcγ receptor binding correlates with functional activity against SARS-CoV-2 variants**

**Table S1: Demographic and clinical description of the wave 1 and wave 2 cohorts**

| Patient ID | Wave | Gender | Age Range | Days from test to sample | Confirmed variant by sequence |
|---|---|---|---|---|---|
| COV004 | 1 | Male | 61-65 | 8 | |
| COV005 | 1 | Male | 36-40 | 15 | |
| COV006 | 1 | Male | 26-30 | 8 | |
| COV020 | 1 | Male | 51-55 | 10 | |
| COV023 | 1 | Male | 26-30 | 12 | |
| COV024 | 1 | Male | 66-70 | 7 | |
| COV026 | 1 | Female | 56-60 | 14 | |
| COV027 | 1 | Male | 51-55 | 10 | |
| COV028 | 1 | Female | 41-45 | 8 | |
| COV030 | 1 | Male | 41-45 | 10 | |
| COV031 | 1 | Female | 41-45 | 17 | |
| COV033 | 1 | Female | 46-50 | 11 | |
| COV034 | 1 | Male | 56-60 | 14 | |
| COV036 | 1 | Male | 41-45 | 9 | |
| COV039 | 1 | Male | 36-40 | 10 | |
| COV040 | 1 | Male | 41-45 | 10 | |
| COV043 | 1 | Male | 51-55 | 10 | |
| COV044 | 1 | Male | 46-50 | 10 | |
| COV045 | 1 | Female | 60-65 | 8 | |
| COV046 | 1 | Male | 41-45 | 9 | |
| COV048 | 1 | Male | 70-75 | 9 | |
| COV049 | 1 | Female | 51-55 | 23 | |
| COV052 | 1 | Female | 56-60 | 12 | |
| COV053 | 1 | Female | 61-65 | 13 | |
| COV055 | 1 | Female | 51-55 | 33 | |
| COV061 | 1 | Male | 61-65 | 13 | |
| COV076 | 1 | Male | 71-75 | 7 | |
| SA-01-0001 | 2 | Female | 26-30 | 12 | |
| SA-01-0005 | 2 | Male | 46-50 | 17 | |
| SA-01-0006 | 2 | Female | 46-50 | 21 | |
| SA-01-0007 | 2 | Female | 46-50 | 16 | |
| SA-01-0011 | 2 | Female | 21-25 | 7 | |
| SA-01-0017 | 2 | Female | 66-70 | 12 | |
| SA-01-0018 | 2 | Male | 61-65 | 29 | |
| SA-01-0022 | 2 | Male | 46-50 | 2 | |
| SA-01-0028 | 2 | Male | 66-70 | 20 | |
| SA-01-0032 | 2 | Female | 61-65 | 14 | |
| SA-01-0033 | 2 | Male | 56-60 | 3 | |
| SA-01-0034 | 2 | Male | 41-45 | 6 | |
| SA-01-0038 | 2 | Male | 46-50 | 13 | |
| SA-01-0042 | 2 | Female | 41-45 | 29 | |
| SA-01-0061 | 2 | Male | 56-60 | 25 | |
| SA-01-0066 | 2 | Male | 61-65 | 2 | |
| SA-01-0068 | 2 | Male | 31-35 | 20 | |
| SA-01-0070 | 2 | Female | 66-70 | 8 | |
| SA-01-0084 | 2 | Female | 55-60 | 2 | |
| SA-01-0088 | 2 | Female | 71-75 | 15 | |
| SA-01-0092 | 2 | Female | 66-70 | 11 | |
